# Pittsburgh plasma p-tau217: classification accuracies for autosomal dominant and sporadic Alzheimer’s disease in the community

**DOI:** 10.1101/2025.05.03.25326526

**Authors:** Anuradha Sehrawat, Xuemei Zeng, Eric E. Abrahamson, Rebecca A. Deek, Alexandra Gogola, M Ilyas Kamboh, Tharick A. Pascoal, Victor L. Villemagne, Oscar L. Lopez, Milos D. Ikonomovic, Beth E. Snitz, Ann D. Cohen, Thomas K. Karikari

## Abstract

**INTRODUCTION:** Most available p-tau217 immunoassays have similar performances. It is unclear if this is due to the use of the same antibody (the “ALZpath antibody”). We established and evaluated a novel p-tau217 assay that employs an alternative antibody, and benchmarked the results against ALZpath-p-tau217.

**METHODS:** Following development and analytical validation of the University of Pittsburgh (“Pitt-p-tau217”) method, clinical verification was performed in three independent cohorts (n=363).

**RESULTS:** Pitt-p-tau217 demonstrated high between-run stability, linearity, and specificity. Clinically, Pitt-p-tau217 differentiated neuropathologically confirmed *PSEN1* mutation carriers from controls with AUC=0.94, and Aβ-PET-positive from Aβ-PET-negative cognitively normal older adults with AUC up to 0.84, equivalent to ALZpath-p-tau217 results. Both Pitt-p-tau217 and ALZpath-p-tau217 were elevated in tau-PET-positive versus tau-PET-negative participants (P=0.06; AUC=0.71 for both). Between-assay correlations were up to 0.93.

**DISCUSSION:** The new Pitt-p-tau217 assay exhibits high and reproducible classification accuracies for identifying individuals with biological evidence of AD, equivalent to the widely used ALZpath-p-tau217.

## 1 INTRODUCTION

The last five years has witnessed a substantial surge in the number of studies evaluating the diagnostic and prognostic utility of plasma biomarkers for Alzheimer’s disease (AD), particularly phosphorylated tau (p-tau)^1–3^. Plasma p-tau181 was the first blood-based p-tau biomarker to be described^4–6^ and was quickly followed by p-tau217^7^, p-tau231^8^ and lately p-tau212^9^. Furthermore, multi-analyte methods employing immunoassay or mass spectrometry approaches that provide concurrent measurement of these analytes are now available^10, 11^. A converging point in the dozens of studies evaluating plasma p-tau is that the levels of these biomarkers are elevated according to the severity of AD pathophysiology while individuals presenting with non-AD neurogenerative diseases show normal levels similar to those of unaffected controls^4–8, 12,13, 14^. These findings highlight the value of plasma p-tau biomarkers for AD prognosis, diagnosis, staging and differential diagnosis.

Recent investigations have suggested that plasma p-tau217 tends to demonstrate superior diagnostic and prognostic accuracies compared with p-tau181 and p-tau231 especially in the preclinical stages^13–15^ while another study reported equivalence of plasma p-tau217 with p-tau212 ^12^. To this end, plasma p-tau217 is poised to be a leading blood-based AD biomarker as it is gradually being applied in therapeutic trials and in referral clinics^15^.

Nonetheless, there are a few notable gaps in the use of plasma p-tau217. Firstly, most of the commercial immunoassays currently available use an identical anti-p-tau217 antibody from ALZpath Inc.; these include methods from leading entities providing *in vitro* diagnostic and laboratory developed tests based on which most of the current literature on plasma p-tau217 has been gathered. Examples of these p-tau217 assays include, but not limited to, those from ALZpath/Quanterix, Roche Diagnostics, Beckman Coulter, NULISA/Alamar Biosciences, SpearBio, among others. Although one could argue that using the same antibody helps streamline results on p-tau217, the reverse argument also holds true – this practice precludes in-depth validation of the performance of p-tau217 toward establishing its accuracies when antibodies from different sources but targeting the same analyte are employed. Secondly, studies in community-population-based cohorts are limited^16–18^, so are investigations focusing on autosomal dominant AD (ADAD)^7, 19^.

In this study, we performed biochemical evaluation of a novel p-tau217 antibody against synthetic and recombinant tau reagents as well as by immunohistochemistry of human AD brain tissue. Next, we applied this antibody to develop a new plasma p-tau217 method, hereafter referred to as Pitt-p-tau217 (given that it originated from the University of Pittsburgh) followed by its technical validation. In clinical validation, we benchmarked the p-tau217 assay performance against that of the widely used commercial p-tau217 method from ALZpath/Quanterix. Finally, we report the assay accuracies in a blood-to-autopsy cohort of ADAD participants as well as in population-based cohorts of older adults who were predominantly without cognitive symptoms but, had received comprehensive brain imaging assessments for amyloid plaque and tau tangle pathologies.

## 2 METHODS

### 2.1 Biochemical characterization of p-tau217 antibody

Sandwich enzyme-linked immunosorbent assays (ELISAs) were used to ascertain the specificity of an anti-p-tau217 rabbit monoclonal antibody (mAb) from Cell Signaling Technology. For each measurement, 80 μl of the rabbit mAb at 0.6 μg/ml or the anti-tau mouse mAb clone Tau5 (epitope: amino acids 210-230 on tau441; BioLegend, #806403) at 1.5 μg/ml, in PBS pH 7.2 (Gibco, #20012027) was added into the well and incubated overnight at 4°C. The well was then blocked with 200 μl PBS/0.1% BSA (Millipore, #81-053-3) for one hour at room temperature, and washed twice with 300 μl PBST (PBS with 0.1% Tween 20, Bio-Rad, #1610781). Subsequently, 50 μl of recombinant tau441 (SignalChem, #T08-54N) or its GSK3beta-phosphorylated variant (p-tau441; SignalChem, #T08-50FN) at concentrations ranging from 0 to 100 ng/ml were added, followed by the addition of 50 μl of PBST with 2% milk (Lab Scientific bioKEMIX, Inc., #978-907-4243) and 50 μl of biotinylated anti-tau mouse mAb, clone Tau12 (directed at tau441 amino acids 6-18; BioLegend, #806502) at 1 μg/ml. Immunocomplex formation proceeded for one hour with gentle shaking at 300 rpm and room temperature. The wells were washed five times with 300 μl PBST. Streptavidin-HRP (Pierce, #21130) was added, and the mixture incubated for one hour at room temperature, followed by five washes with 300 μl PBST. The TMB substrate (Thermo Scientific, #34022) was added and allowed to incubate for 30 minutes before stopping the reaction with 100 μl of stop solution (Thermo Scientific, #N600). A spectrophotometer (Type, Model, Source) was used to determine the optical density (OD) at 450 nm with subtraction of the background OD at 550 nm.

### 2.2 Immunohistochemistry

Autopsy brain tissue collection and analyses were approved by the University of Pittsburgh Committee for Oversight of Research and Clinical Training Involving Decedents. For immunohistochemistry, brain tissue used was from a non-Hispanic White participant (aged between 60-70 years) neuropathologically diagnosed having a high degree of AD neuropathological change (Braak stage VI, high likelihood of AD by the NIA-RI criteria^20, 21^) with sparse neocortical alpha-synuclein pathology and no TAR DNA binding protein 43 (TDP-43) pathology. Post-mortem delay was two hours. Blocks of tissue from the middle temporal gyrus and from the hippocampus were obtained at autopsy and immediately immersed in ice-cold 4% paraformaldehyde made in 0.1% sodium phosphate buffer (PB) for 48 hours. Tissue blocks were then sequentially immersed in 15% sucrose and 30% sucrose both made in PB (24 hours each), frozen, and cut into 40 µm thick sections using a freezing, sliding microtome. Tissue sections were then immersed in cryoprotectant solution (1% polyvinylpyrrolidone, 30% sucrose, and 30% ethylene glycol made in PB) and stored at -20 degrees Celsius until used for experiments. Before use, tissue sections were extensively rinsed in PB to remove the cryoprotectant solution. Immunohistochemistry was performed on free-floating sections as described previously^22^ using the VECTASTAIN Elite ABC-HRP kit (Vector Laboratories, Burlingame, CA, #PK-6100) with nickel enhanced 30,3-diaminobenzidine tetra-hydrochloride (DAB, Sigma, D8001) as the chromogen. The primary antibody was a rabbit monoclonal IgG (clone E9Y4S, diluted 1:1,000) directed against the tau phospho-epitope at threonine 217 (Cell Signaling Technologies, #51625BF). According to the manufacturer, this antibody labels a band at the expected molecular weight for tau when assessed by Western blot, and this band is absent in tau-knockout mice.

### 2.3 Plasma p-tau217 immunoassay development and technical validation

Assay development, validation and clinical studies were performed at the Biofluid Biomarker Laboratory, Department of Psychiatry, School of Medicine, University of Pittsburgh, Pittsburgh, PA, USA. A two-step ultra-sensitive p-tau217 immunoassay was developed on the Simoa HD-X instrument (Quanterix, MA, USA).

For capture, the rabbit p-tau217 mAb was coupled to paramagnetic beads (Quanterix, #103207). Biotin-conjugated mouse monoclonal antibody raised against the N-terminal region of tau (Tau12; BioLegend, #SIG-39416) was used for detection. *In vitro* phosphorylated recombinant full-length tau441 (SignalChem, #T08-50FN-50) was used as the assay calibrator. Plasma samples and calibrators were diluted with assay diluent (sample diluent, Quanterix, #104101). Optimal required dilution of samples was established as 1:4, since at this dilution measured samples gave signals around the middle of the standard curve.

Analytical validation focused on dilution linearity, competitive binding, day-to-day stability, and spike recovery was done as described previously^23^. Sensitivity (lower limit of quantification; LLOQ) was calculated from a three-point standard curve using recombinant p-tau441 (0-96 pg/mL; in duplicates). These curves were generated on two distinct HDX instruments, utilizing different lots of antibody-conjugated paramagnetic beads, each prepared on separate days. For assay specificity, a p-tau217-containing antigen (synthetic peptide sourced from Sino Biologicals US Inc.) at varying concentrations of 0.1-200 μg/ml was used for competitive binding with endogenous p-tau217 in plasma samples. In brief, p-tau217-containing antigen peptide concentrations ranging from 0.1 to 200 µg/ml were added to plasma samples, followed by 4X dilutions with assay diluent, and then analyzed using the Pitt-p-tau217 assay.

### 2.4 Biomarker clinical studies

In the clinical studies, p-tau217 was measured in 1:4 dilution of plasma samples by scientists blinded to participant information. Plasma samples were analyzed in singlicates while signal variations within and between analytical runs were monitored using three internal quality controls (iQC) samples measured in duplicates at the start and the end of each technical run.

For comparative purposes, plasma p-tau217 was also measured with a commercial method from Quanterix, Inc. (ALZpath-p-tau217 assay kit #103714) using the Simoa HD-X platform for a selection of cohorts.

### 2.5 Cohorts

#### ADAD study

This cohort included plasma samples from individuals who underwent brain autopsies conducted by the Neuropathology Core of the University of Pittsburgh Alzheimer’s Disease Research Center (ADRC). The neuropathological diagnosis was determined as described previously, using the CERAD criteria^24^, Braak neurofibrillary tangle staging^25^, and Thal amyloid-β phases^26^. This resulted in a composite ABC score for each case according to the NIA-AA criteria^20^. The clinical evaluation for diagnosing Alzheimer’s disease dementia adhered to established criteria. Subgroups were identified with neurochemically defined AD and AD dementia. Of these, some had sporadic AD and others with ADAD. Five sporadic AD and two ADAD participants underwent [^11^C]PiB positron emission tomography (PET) imaging for Aβ and later came to autopsy. Five out of six cases classified as ADAD were confirmed to have a defined *PSEN1* mutation through genetic testing. Control participants, matched in age and sex, without AD profiles as determined by clinical evaluation, were selected from the ADRC for comparison.

#### MYHAT-NI (Monongahela-Youghiogheny Healthy Aging Team-Neuroimaging) study

The MYHAT-NI is a sub-study of the parent MYHAT study which is a population-based research cohort that focuses on selected communities from a Rust Belt region of southwestern Pennsylvania, USA. The participants were selected through age-stratified random sampling from the publicly available voter registration lists during two time periods: 2006–2008 and 2016– 2019. The eligibility criteria upon enrollment encompassed: (1) being aged 65 years or older, (2) residing in a specified town, (3) not living in long-term care facilities, (4) possessing adequate hearing and vision for completing neuropsychological assessments, and (5) be able to make decisions. Participants were categorized as cognitively normal if they scored 0 on the Clinical Dementia Rating (CDR), classified as mildly cognitively impaired with a CDR of 0.5, and diagnosed with dementia if their CDR was ≥1. Aβ PET was assessed using [^11^C]PiB PiB and tau PET employed [^18^F]MK-6240.

#### Human Connectome Project (HCP) cohort

The HCP cohort is an ongoing longitudinal, community-based study in Pittsburgh PA, USA, examining brain structural and functional connectivity among individuals aged 56–79 years, including both cognitively normal and cognitively impaired participants. The HCP study employs the Pittsburgh ADRC neuropsychological test battery assessments (including the Montreal Cognitive Assessment (MoCA), verbal fluency, a 30-item visual naming test, Trail making verbal free recall and the Rey-Osterreith Complex Figure), along with brain structural and functional imaging techniques such as functional MRI and magnetoencephalography to evaluate brain connectivity. Additionally, [^11^C]PiB PET imaging was conducted *in vivo* to examine brain Aβ pathology. Participants were classified into normal, impaired without complaints, subjective cognitive complaints, mild cognitive impairment (MCI, both amnestic and non-amnestic), and AD using the ADRC classification scheme ^27^.

In the MYHAT-NI and HCP cohorts, we employed previously documented approaches to determine Aβ PET positivity standardized uptake value ratio (SUVR) >1.346^28, 29^. Aβ PET SUVR values were converted to the Centiloid scale^30^ using established cutoff values: Aβ-negative (CL<15), low-burden Aβ (CL15-25), and Aβ-positive (CL>25).

### 2.6 Ethical clearance

The University of Pittsburgh Institutional Review Board approved the study procedures, and written informed consent was obtained from all participants; The reference numbers for the study approvals are as follows: ADAD study, MOD19110245-023; MYHAT-NI, STUDY19020264; and HCP, STUDY19100015C.

### 2.7 Statistical analyses

Continuous data are shown as median and interquartile range (IQR). Categorical variables are reported as counts (%). Group differences in continuous variables were assessed using the Wilcoxon Rank Sum test (for two categories) or the Kruskal-Wallis test with Dunn’s multiple comparisons (for three or more groups), with p-values adjusted using Bonferroni correction. Likewise, group differences in categorical variables were assessed using Fisher’s exact test.

The overall association between biomarker pairs within each study was reported using Spearman correlation. Multivariable logistic regression models were built to assess the performance of each p-tau217 assay in predicting Aβ-PET and Tau PET status. The primary predictor of these models was the biomarker of interest. Additionally, the models were adjusted for age, sex, and *APOE* ε4 carrier status as covariates. Models were fit for each biomarker and each study separately. Performance was assessed using three-fold cross validation, in which the data is trained on two-thirds of the data and tested on the remaining one-third, that was not used in model building. This process was repeated three times, once for each of the “left out” folds. For MYHAT-NI and HCP cohorts diagnostic potential was examined using receiver operating curves and area under the curve (AUC-ROC). The reported AUC is the average of the test set AUCs across folds. The AUC from different biomarkers were compared by examining the overlap in their confidence intervals, estimated using the method proposed previously^31^. Python was used to measure AUC-ROC for ADAD study. Throughout the analyses, statistical significance was set at p<0.05. Three data points, one from the ADAD study and two from the HCP cohort, that fell outside the calibration curve range were excluded from the final analysis. Statistical analyses were performed with Python (version 3.12) and R (version 4.4.2). Plots were generated with Python, R or GraphPad Prism (version 10.1.1).

## 3 RESULTS

### 3.1 Antibody specificity to the p-tau217 site

We employed sandwich ELISA to evaluate binding profiles of serial dilutions (0-100 ng/ml) of the p-tau217 rabbit mAb to recombinant full-length tau441 protein either non-phosphorylated or phosphorylated jointly at threonine-217^32^ and multiple other sites including p-tau181^4^ and p-tau231^8^. The Tau5 antibody, which was raised against the non-phosphorylated tau peptide aa210-230 spanning the threonine-217 site, was used as a control. The p-tau217 antibody exhibited no recognition to non-phosphorylated tau441 (Figure 1A) but showed dose-dependent binding to the phosphorylated version known to contain the p-tau217 site (Figure 1B). Tau5, on the other hand, did bind non-selectively to both the phosphorylated and non-phosphorylated tau441 recombinant proteins (Figure 1A-B).

**Figure 1.**
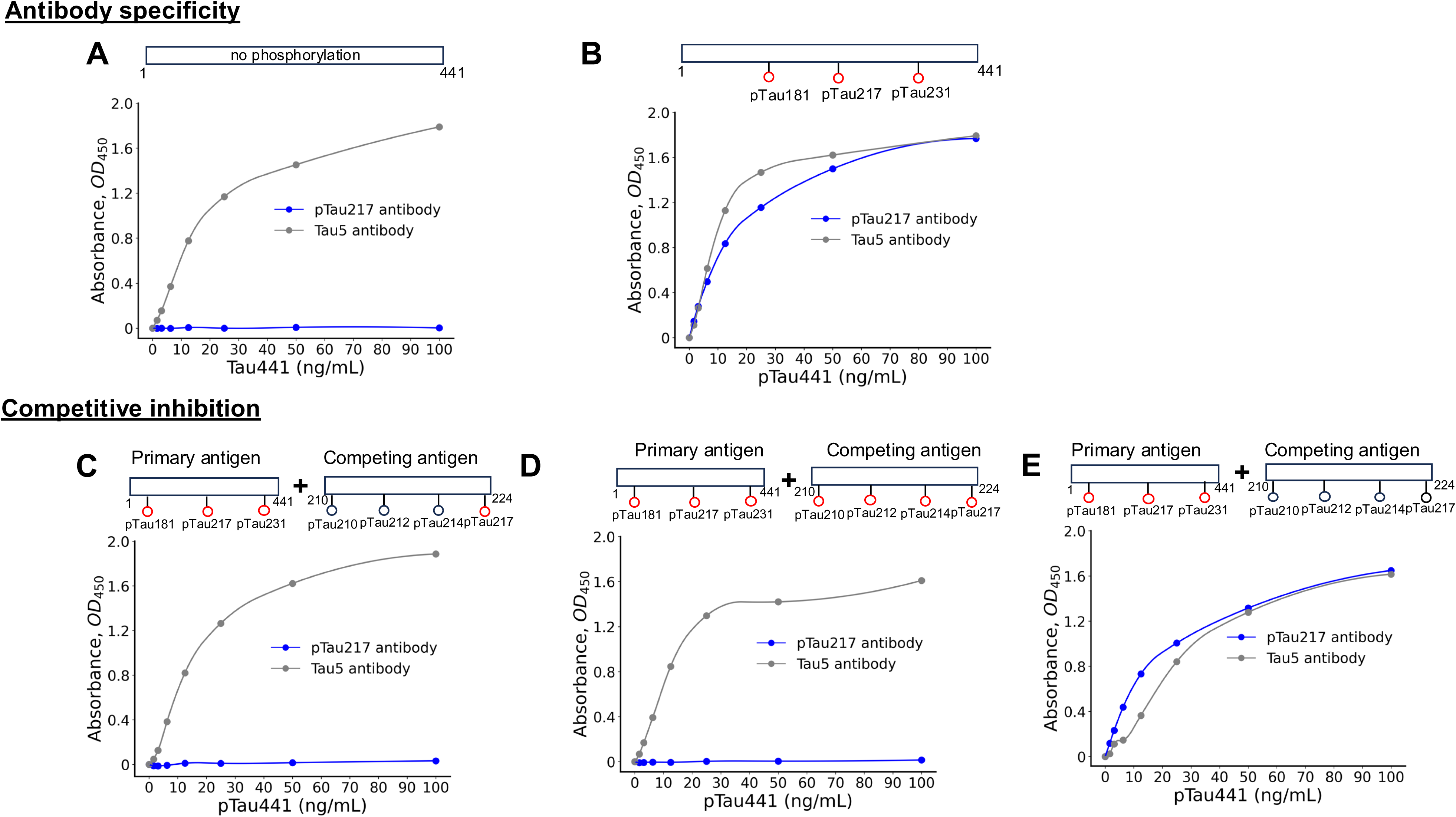
Biochemical characterization of the p-tau217-specific rabbit monoclonal antibody. Each panel displays the results of sandwich ELISAs, utilizing either the p-tau217-specific rabbit monoclonal antibody or the anti-tau mouse monoclonal antibody, clone Tau5, for capture. Detection was performed using a biotinylated anti-tau mouse monoclonal antibody, clone Tau12 – the same antibody used for detection in the eventual Pittsburgh p-tau217 assay. The assays tested varying concentrations of recombinant tau441 (non-phosphorylated tau441 (2N4R) isoform) or its GSK3beta-phosphorylated variant (p-tau441) either alone (A and B) or against a set concentration (0.1 μg/ml) of a synthetic peptide (C-E). These peptides, corresponding to tau441 amino acids 210 to 224 (SRTPSLPTPPTREPK), were linked to an N-terminal cysteine via a peptide bond and phosphorylated at the residues specified in the illustrations in the figure. Panels (A-B) depict the binding profiles of the p-tau217 and Tau5 antibodies to recombinant non-phosphorylated tau441 (A) and phosphorylated tau441 (B). Panels (C-E) show the binding profiles of the p-tau217 and Tau5 antibodies to phosphorylated tau441 in the presence of synthetic peptides phosphorylated exclusively at threonine-217 (C), peptides phosphorylated jointly at serine/threonine 210, 212, 214, and 217 (D), and non-phosphorylated peptides (E).

Next, we asked if a p-tau217-containing synthetic peptide can competitively inhibit the binding of the p-tau217 antibody to recombinant phosphorylated tau441. We generated phosphorylation variants of an N-terminal cysteine-labelled synthetic tau210-224 peptide which covers the threonine-217 site. In the presence of synthetic peptides phosphorylated exclusively at threonine-217 or jointly at serine/threonine 210, 212, 214, and 217, binding of the p-tau217 antibody was attenuated (Figure 1C-D) but not when the synthetic peptide added was not phosphorylated at any site (Figure 1E). None of these synthetic peptides affected the binding of Tau5 to recombinant phosphorylated tau441 (Figure 1C-E).

Together, these results show that the p-tau217 antibody used in this study is specific to phosphorylation at threonine-217, and its binding is unaffected by the phosphorylation status of adjacent serine or threonine residues.

### 3.2 P-tau217 antibody stains neurofibrillary tau pathology in human AD brain tissue

We examined the staining pattern of the p-tau217 antibody. Immunohistochemistry was performed on free-floating tissue sections from the middle temporal gyrus (BA21) (Figure 2) and hippocampus (Figure 3) from an end-stage AD case (Braak VI; age at death, 60-70 years).

**Figure 2.**
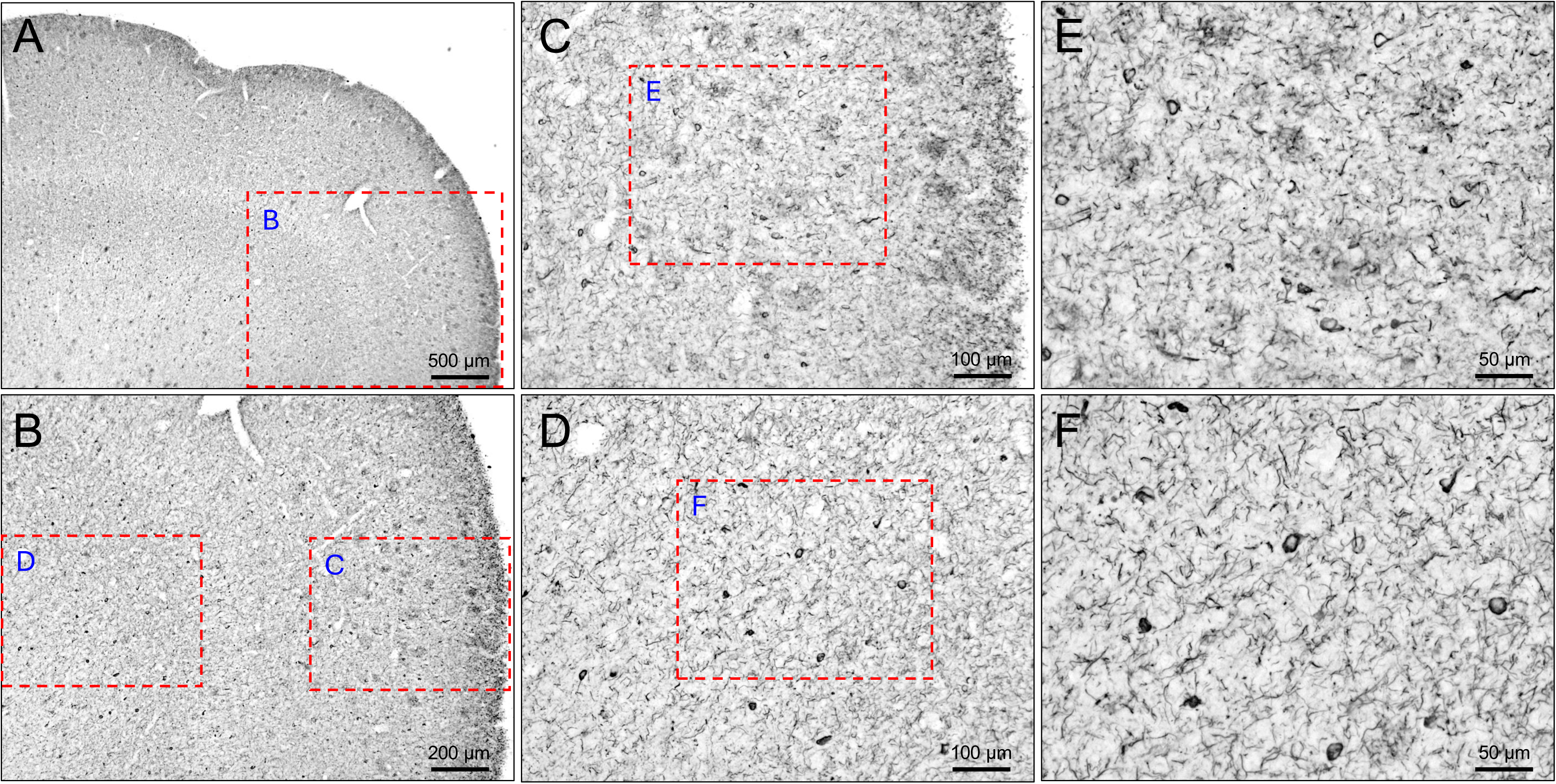
P-tau217 antibody stains neurofibrillary tangles in middle temporal gyrus region of human brain tissue. Section of middle temporal gyrus from an AD case neuropathologically diagnosed as Braak stage VI, immunohistochemically stained with p-tau217 antibody. A dense network of immunoreactive structures is present throughout the cortical laminae illustrated at low magnification in (A). The dashed red box in (A) delineates the area shown at higher magnification in panel (B). The dashed red boxes in (B) delineate areas illustrated at higher magnifications in panels (C, superficial lamina) and (D, deep lamina). The dashed red boxes in (C) and (D) delineate areas illustrated at higher magnifications in panels (E) and (F), respectively. Immunoreactive clusters of dystrophic neurites in neuritic plaques are most numerous in superficial lamina, while immunoreactive neurofibrillary tangles and neuropil threads are numerous throughout the cortical laminae. Scale bars = 500 µm (A), 200 µm (B), 100 µm (C, D), and 50 µm (E, F).

**Figure 3.**
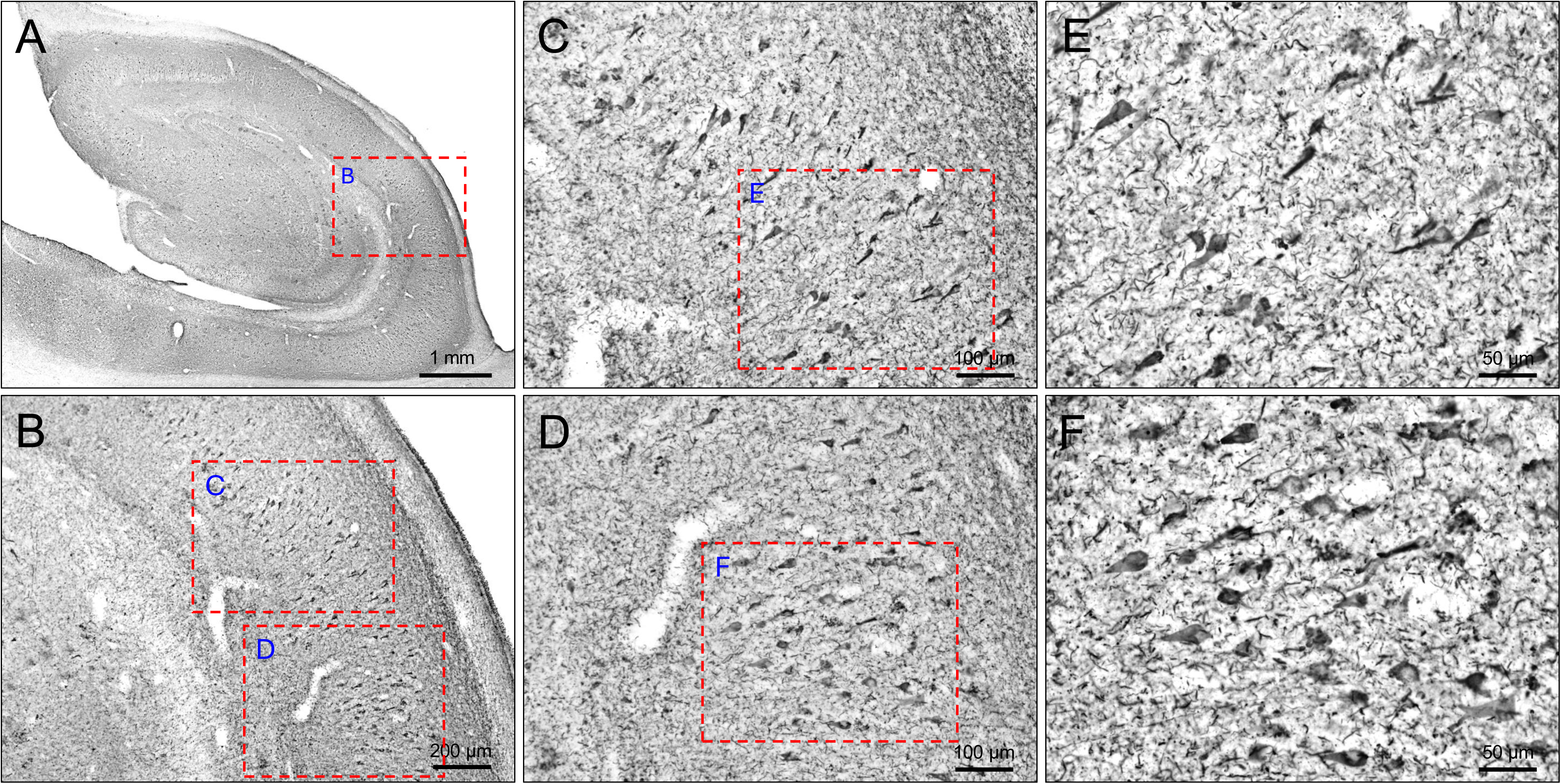
P-tau217 antibody stains neurofibrillary tangles in hippocampal region of human brain tissue. Section of hippocampus from an AD case neuropathologically diagnosed as Braak stage VI, immunohistochemically stained with p-tau217 antibody. Numerous immunoreactive neurofibrillary tangles are present in the CA fields, immunoreactive dystrophic neurites in neuritic plaques are abundant in the dentate gyrus stratum moleculare, and immunoreactive neuropil threads are distributed throughout the CA fields and dentate gyrus illustrated in a low magnification composite in panel (A). The dashed red box in (A) delineates the area (field CA1 and dentate stratum moleculare) shown at higher magnification in panel (B). The dashed red boxes in (B) delineate areas illustrated at higher magnifications in panels (C) and (D). The dashed red boxes in (C) and (D) delineate areas illustrated at higher magnifications in panels (E) and (F), respectively. Immunoreactive neurofibrillary tangles embedded in a mesh of neuropil threads are the most prominent features of the CA1 area. Many immunoreactive neurons had a classic flame-shaped tangle morphology, while others appeared to contain more homogeneous immunoreaction and morphology of intracellular tangles.

In sections of middle temporal gyrus (Figure 2), p-tau217 immunoreactivity was detected in neurofibrillary tangles (NFT), in clusters (40-50 µm in diameter) of dystrophic neurites indicative of neuritic plaques, and in threads distributed throughout the neuropil. Immunoreactive clusters of dystrophic neurites were most numerous in superficial cortical lamina (Figure 2 A-C,E) while neurofibrillary tangles were more numerous in deeper cortical laminae (Figure 2A,B,D,F). Overall distribution and density of immunoreactivity exhibited a moderately laminar pattern, being higher in the supragranular lamina and infragranular lamina than lamina IV.

In sections of the hippocampus (Figure 3), p-tau217 immunoreactivity was detected in NFT, dystrophic neurites/neuritic plaques and neuropil threads. NFT were most numerous in hippocampal CA1 field (Figure 3 A-F) and were also present in other CA fields and in the dentate stratum granulosum. Dystrophic neurites/neuritic plaques were most numerous in the dentate gyrus stratum moleculare and the hilus and immunoreactive neuropil threads were observed throughout the hippocampus and dentate gyrus and were most numerous in CA1 (Figure 3).

### 3.3 Development and validation of the Pittsburgh p-tau217 assay

We developed a Simoa-based immunoassay to detect p-tau217 in plasma. This involved utilizing the new rabbit antibody validated as described above for capture alongside the mouse monoclonal antibody Tau12, which targets the N-terminal tau 6-18 epitope.

Following assay optimization, technical performance was evaluated according to guidelines provided by an international consortium of clinical chemists^23^. The p-tau217 assay exhibited robust linear dilution properties, with signals decreasing proportionally when measured at two-, four-, or eight-fold dilutions (Figure 4A). These results were consistent for three different pooled plasma samples of variable starting concentrations.

**Figure 4.**
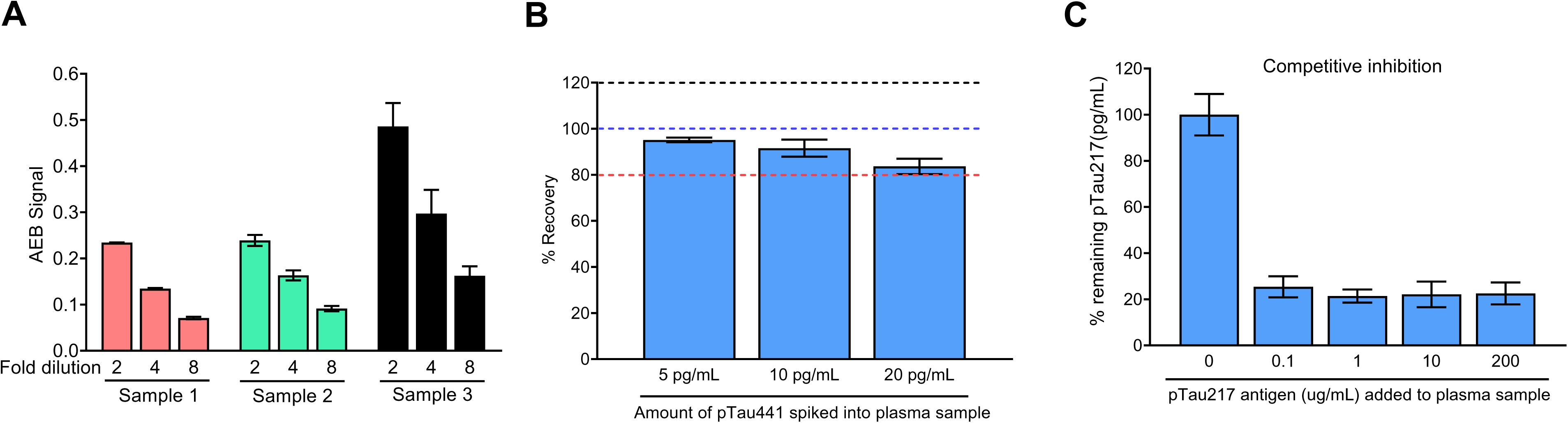
Technical validation of the newly developed novel plasma p-tau217 assay. Dilution linearity of the p-tau217 assay in pooled plasma samples (A). For each matrix, the plots show the measured AEB signal in three unique samples with variable levels of the biomarker. Three equal-volume aliquots of each sample were prepared and measured diluted 2-, 4-, or 8-fold with the assay diluent. Samples were run in duplicates. (B) Percentage recovery of the p-tau217 in pooled plasma sample spiked with assay calibrator of concentrations 5,10 and 20 pg/ml. Samples at each concentration were assayed in duplicates. The dashed line represents the acceptable recovery range between 80-120%. (C) Assay specificity of the newly developed plasma p-tau217 assay by competitive inhibition. The plot shows the results of competitive inhibition experiments of p-tau217 concentrations in a pooled plasma sample that was either untreated or inhibited with the addition of different amounts (range: 0.1-200 ug/ml) of a synthetic peptide bearing phosphorylation at the threonine-217 site. Data points represent mean concentration of plasma p-tau217, measured in duplicates in two independent experiment runs and error bars represent ± SD. AEB, average enzyme per bead.

In spike recovery, the exogenous addition of recombinant p-tau441 at three different concentrations (5, 10, and 20 pg/ml) to the test plasma sample was performed. High efficiency in recovery of the expected signal (83.6 to 95.1%) was observed (Figure 4B), indicating a lack of significant matrix interference. The calculated average within-run imprecision was recorded at 3.6%-8.8% and the between-run imprecision at 4%-16.8% when three independent samples were tested across up to five separate analytical runs (Supplemental Figure 1 A and B). The lower limit of quantification for the assay (LLOQ) was estimated to be 0.321 pg/mL.

Further examination of assay robustness was conducted by running a nine-point standard curve (recombinant p-tau441; 0–96Lpg/ml) on two distinct HDX instruments and using different lots of antibody-conjugated paramagnetic beads prepared on separate days. Similar signals were observed at each data point, demonstrating high precision across reagent batches (Supplemental Figure 2).

### 3.4 Assay specificity

To assess assay specificity, plasma samples were tested for p-tau217 levels in the presence and absence of various concentrations (0.1-200 μg/mL) of a p-tau217-positive antigen-the same synthetic peptide used in Figure 1C. The results of this competitive inhibition experiment showed that ∼80% of the p-tau217 signal was masked in the presence of as low as 0.1 μg/mL of the synthetic peptide (Figure 4C). These results show that the assay signal was specific to p-tau217.

### 3.5 Participants

#### We included a total of 363 participants from three independent cohorts

The ADAD cohort (n=21) included autopsy-verified individuals with sporadic AD (n=8) and ADAD due to *PSEN1* mutations (n=6) as well as non-AD controls (n=7). All cases classified as ADAD had the *PSEN1* mutation confirmed by genetic testing, except for one case. The autopsy-confirmed participants, encompassing both sporadic and ADAD, had clinical history of subjective dementia. Non-AD controls were cognitively unimpaired individuals that had no or low levels of AD neuropathologic changes at autopsy.

The MYHAT-NI cohort included 176 plasma samples from 113 participants (Table 1): 63 (55.8%) provided plasma samples at both the baseline and 2-year visits. At baseline (n=113; median age: 76.0 years [IQR: 72.0–80.0]), 54% were female, 95% self-identified as non-Hispanic White (NHW) and 28 (25%) participants) were Aβ-PET positive (A+). At baseline, 39 (35%) were tau PET positive (T+). Majority of the participants (88%) had normal cognition (CDR=0) at both visits. The proportion of *APOE* ε4 carriers was higher in the A+ compared to the Aβ-PET negative (A-) groups at both baseline and 2-year visits, with p values of <0.001 and 0.014, respectively. Additionally, being A+ was significantly associated with higher tau PET uptake, with p-values of <0.001 for baseline and 0.001 for the 2-year visit.

**Table 1:** Demographic characteristics of the MYHAT-NI and HCP cohorts.

|  | MYHAT-NI Cohort |  |  |  |  |  |  |  |  | HCP Cohort |  |  |  |
| --- | --- | --- | --- | --- | --- | --- | --- | --- | --- | --- | --- | --- | --- |
|  | Total | Baseline |  |  |  | Year 2 |  |  |  |  |  |  |  |
|  |  | Total | Aβ PET- | Aβ PET+ | p-value<sup>a</sup> | Total | Aβ PET- | Aβ PET+ | p-value<sup>a</sup> | Total | Aβ PET- | Aβ PET+ | p-value<sup>a</sup> |
| **n** | **176** | **113** | **85** | **28** |  | **63** | **42** | **21** |  | **229** | **195** | **34** |  |
| **Age (years)** | 76.0<br>[72.0,81.0] | 76.0<br>[72.0,80.0] | 75.0<br>[71.0,79.3] | 79.0<br>[73.5,82.5] | 0.055 | 77.0<br>[724.0,81.5] | 76.0<br>[74.0,81.0] | 78.0<br>[73.8,82.3] | 0.884 | 62.0<br>[56.0,70.0] | 60.0<br>[55.5,67.0] | 72.0<br>[68.2,78.8] | <0.001 |
| **Sex, n (%)** |  |  |  |  | 0.275 |  |  |  | 0.109 |  |  |  | 0.003 |
| Female | 93 (53.8) | 61 (54.0) | 43 (50.6) | 18 (64.3) |  | 32 (50.8) | 18 (42.9) | 14 (66.7) |  | 148 (64.6) | 134 (68.7) | 14 (41.2) |  |
| Male | 83 (47.2) | 52 (46.0) | 42 (49.4) | 10 (35.7) |  | 31 (49.2) | 24 (57.1) | 7 (33.3) |  | 81 (35.4) | 61 (31.3) | 20 (58.8) |  |
| **Race, n (%)** |  |  |  |  | 1.000 |  |  |  | 1.000 |  |  |  | <0.001 |
| Black/African American | 9 (5.1) | 6 (5.3) | 5 (5.9) | 1 (3.6) |  | 3 (4.8) | 2 (4.8) | 1 (4.8) |  | 120 (52.4) | 112 (57.4) | 8 (23.5) |  |
| White | 167 (94.9) | 107 (94.7) | 80 (94.1) | 27 (96.4) |  | 60 (95.2) | 40 (95.2) | 20 (95.2) |  | 105 (45.9) | 79 (40.5) | 26 (76.5) |  |
| Asian |  |  |  |  |  |  |  |  |  | 3 (1.3) | 3 (1.5) |  |  |
| Other |  |  |  |  |  |  |  |  |  | 1 (0.4) | 1 (0.5) |  |  |
| **APOE ε4 allele<sup>b</sup>, n (%)** |  |  |  |  | <0.001 |  |  |  | 0.014 |  |  |  | 0.157 |
| Absent | 146 (83.0) | 95 (84.1) | 79 (92.9) | 16 (57.1) |  | 51 (81.0) | 38 (90.5) | 13 (61.9) |  | 159 (69.4) | 139 (71.3) | 20 (58.8) |  |
| Present | 30 (17.0) | 18 (15.9) | 6 (7.1) | 12 (42.9) |  | 12 (19.0) | 4 (9.5) | 8 (38.1) |  | 69 (30.1) | 55 (28.2) | 14 (41.2) |  |
| **CDR Score<sup>c</sup> n (%)** |  |  |  |  | 0.015 |  |  |  | 0.033 |  |  |  |  |
| 0.0 | 156 (88.6) | 102 (90.3) | 80 (94.1) | 22 (78.6) |  | 54 (85.7) | 39 (92.9) | 15 (71.4) |  |  |  |  |  |
| 0.5 | 15 (8.5) | 10 (8.8) | 4 (4.7) | 6 (21.4) |  | 5 (7.9) | 1 (2.4) | 4 (19.0) |  |  |  |  |  |
| **Tau PET<sup>d</sup>, n (%)** |  |  |  |  | <0.001 |  |  |  | 0.001 |  |  |  |  |
| Negative | 116 (65.9) | 74 (65.5) | 65 (76.5) | 9 (32.1) |  | 42 (66.7) | 34 (81.0) | 8 (38.1) |  |  |  |  |  |
| Positive | 60 (34.1) | 39 (34.5) | 20 (23.5) | 19 (67.9) |  | 21 (33.3) | 8 (19.0) | 13 (61.9) |  |  |  |  |  |
| **Clinical diagnosis<sup>e</sup>, n (%)** |  |  |  |  |  |  |  |  |  |  |  |  | 0.002<sup>f</sup> |
| Control |  |  |  |  |  |  |  |  |  | 161 (70.3) | 143 (73.3) | 18 (52.9) |  |
| MCI |  |  |  |  |  |  |  |  |  | 52 (22.7) | 43 (22.1) | 9 (26.5) |  |
| AD dementia |  |  |  |  |  |  |  |  |  | 5 (2.2) | 2 (1.0) | 3 (8.8) |  |
| Unknown |  |  |  |  |  |  |  |  |  | 11 (4.8) | 7 (3.6) | 4 (11.8) |  |

The median and interquartile range (IQR) are reported for continuous variables. Frequencies and percentages are shown for categorical variables. <sup>a</sup>*P*-values were calculated using the Wilcoxon rank-sum test for a continuous variable and Fisher's exact test for a categorical variable, respectively. In MYHAT-NI cohort: <sup>c</sup>One participant at baseline and 4 at the 2-year visit missed CDR assessment. <sup>d</sup>[<sup>18</sup>F]AV-1451 PET, Mean SUVR $\geq 1.18$ as positive. In HCP cohort: . <sup>b</sup>One Aβ PET- participant missed APOE ε4 allele information. <sup>e</sup>Clinical diagnosis of participants was done by ADRC classification scheme. <sup>f</sup>Unknown wasn't included in statistical inference. Racial identity was self-reported.
Note that the MYHAT-NI cohort consists of predominantly control participants, hence the lack of entries under "Clinical diagnosis."

We included 229 individuals from the HCP cohort (Table 1), of which 34 (15%) were A+ and 195 (85%) were A-. Among the A+ participants (median age = 72 years [IQR: 68.2–78.8]), 14 (41%) were *APOE* ε4 carriers, 14 (41%) were females, and 26 (76.5%) were self-identified as White. In comparison, among the A-participants (median age: 60 years, IQR: 55.5–67.0), 55 (28%) carried the *APOE* ε4 allele, 134 (69%) were female, and the racial composition included 79 (40.5%) self-identified as non-Hispanic White and 112 (57.4%) as African American. A+ status was significantly associated with older age (p < 0.001), a higher likelihood of being male (p = 0.003), being white (<0.001), and having cognitive impairment (MCI or AD; p = 0.002).

### 3.7 Comparison of the Pittsburgh p-tau217 assay with a commercially available method

In the ADAD cohort, plasma p-tau217 levels measured using the new Pittsburgh assay were highly correlated with results from the ALZpath p-tau217 assay in the entire cohort (ρ=0.93, p<0.0001; Figure 5A). Within diagnostic groups, the correlations were: non-AD controls (ρ=0.60, p=0.24), sporadic AD (ρ=0.88, p<0.05), and ADAD (ρ=0.94, p=0.017; Supplemental Figure 3), although this led to much reduced sample sizes.

**Figure 5.**
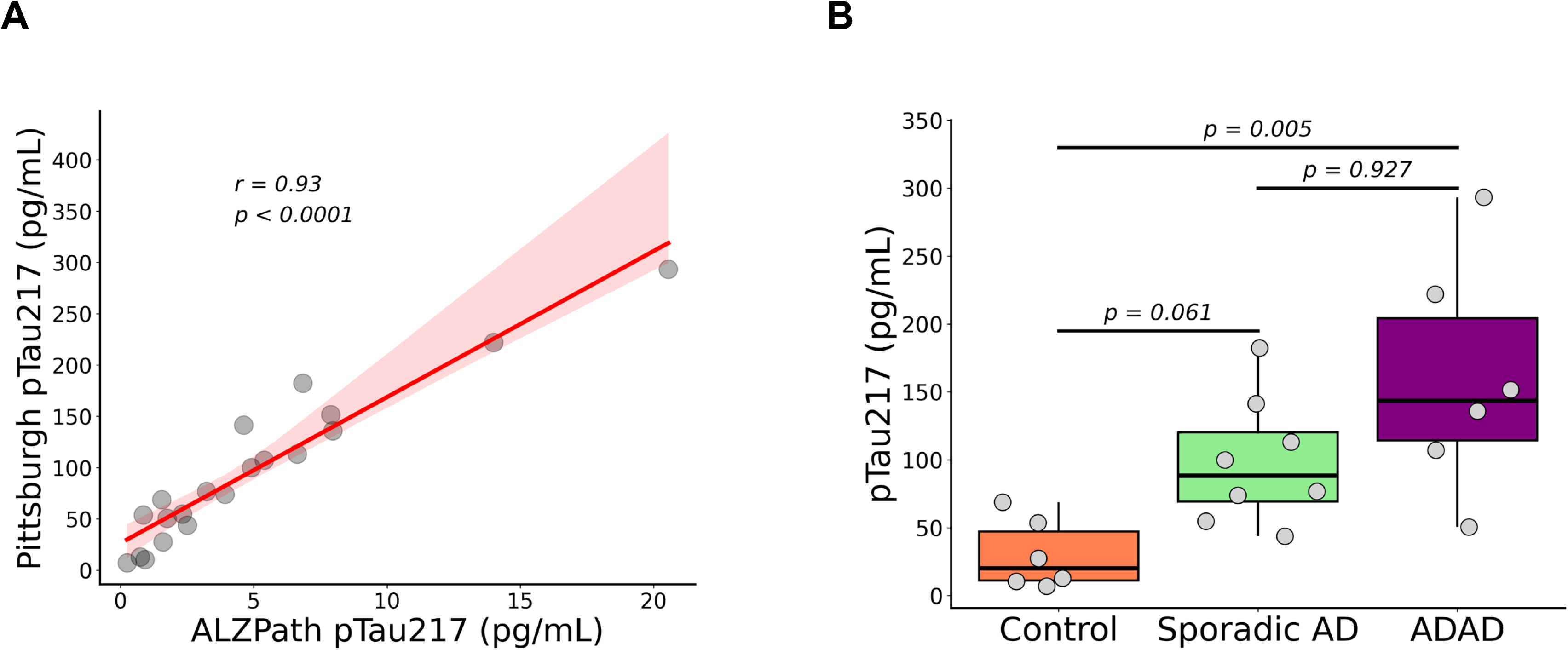
Plasma p-tau217 performance in distinguishing sporadic AD and ADAD from non-AD controls. (A) Correlations between Pitt and ALZpath p-tau217 concentration. The correlation coefficient was calculated using Spearman rank-based correlation. The red line indicates the least square regression line. (B) Boxplot distributions of the p-tau217 levels in ADAD participants with pathogenic *PSEN1* mutations, sporadic AD participants, and control. P-values were derived from post-hoc tests following Kruskal-Wallis, with Bonferroni corrections for multiple comparisons.

Next, we analyzed correlation of the Pittsburgh vs. ALZpath p-tau217 assays in the MYHAT-NI and HCP cohorts. A strong significant correlation was evident across the entire cohort in the MYHAT-NI cohort at baseline (ρ=0.64, p=2.2e-16) and at the two-year follow-up visit (ρ=0.73, p=2.2e-16), as well as in the HCP cohort (ρ=0.3, p=6.7e-06) (Supplemental Figure 4A). Among cognitively normal participants, a similarly strong significant correlation was found in the MYHAT-NI cohort at baseline (ρ=0.62, p=2.2e-16) and at the two-year follow-up (ρ=0.74, p=2.8e-09), as well as in the HCP cohort (ρ=0.37, p=0.00023) (Supplemental Figure 4B).

### 3.6 Discriminative accuracy for sporadic AD and ADAD from control

In the cohort of neuropathologically-confirmed participants, the Pittsburgh p-tau217 assay levels demonstrated stepwise increases from non-AD controls to sporadic AD and then ADAD (Figure 5). In effect, the fold increases in p-tau217 levels were highest in the ADAD group (5.3-fold) followed by the sporadic AD (3.3-fold) versus the non-AD control participants (p=0.005; Figure 5B). The AUC to differentiate ADAD from controls was 94% which was equivalent to 100% for ALZpath.

### 3.8 Accuracy to identify A**β**+ older adults in two community cohorts

In the MYHAT-NI cohort at baseline, plasma p-tau217 levels were 1.8 times higher in Aβ-PET-positive older adults compared with Aβ-PET-negative individuals (Figure 6A) and separated these groups with an AUC of 0.86 (95% CI=0.77-0.94) (Figure 6C). At the two-year visit, this separation was maintained with an AUC of 0.73 (95% CI = 0.61-0.85) (Figure 6C). Similarly, results from the HCP cohort revealed that plasma p-tau217 levels were elevated 1.5-fold in Aβ+ compared with Aβ-individuals (p =0.006; Supplementary Figure 5A), and the two groups were separated with an AUC of 0.86 (95% CI=0.81-0.92) (Figure 6C).

**Figure 6.**
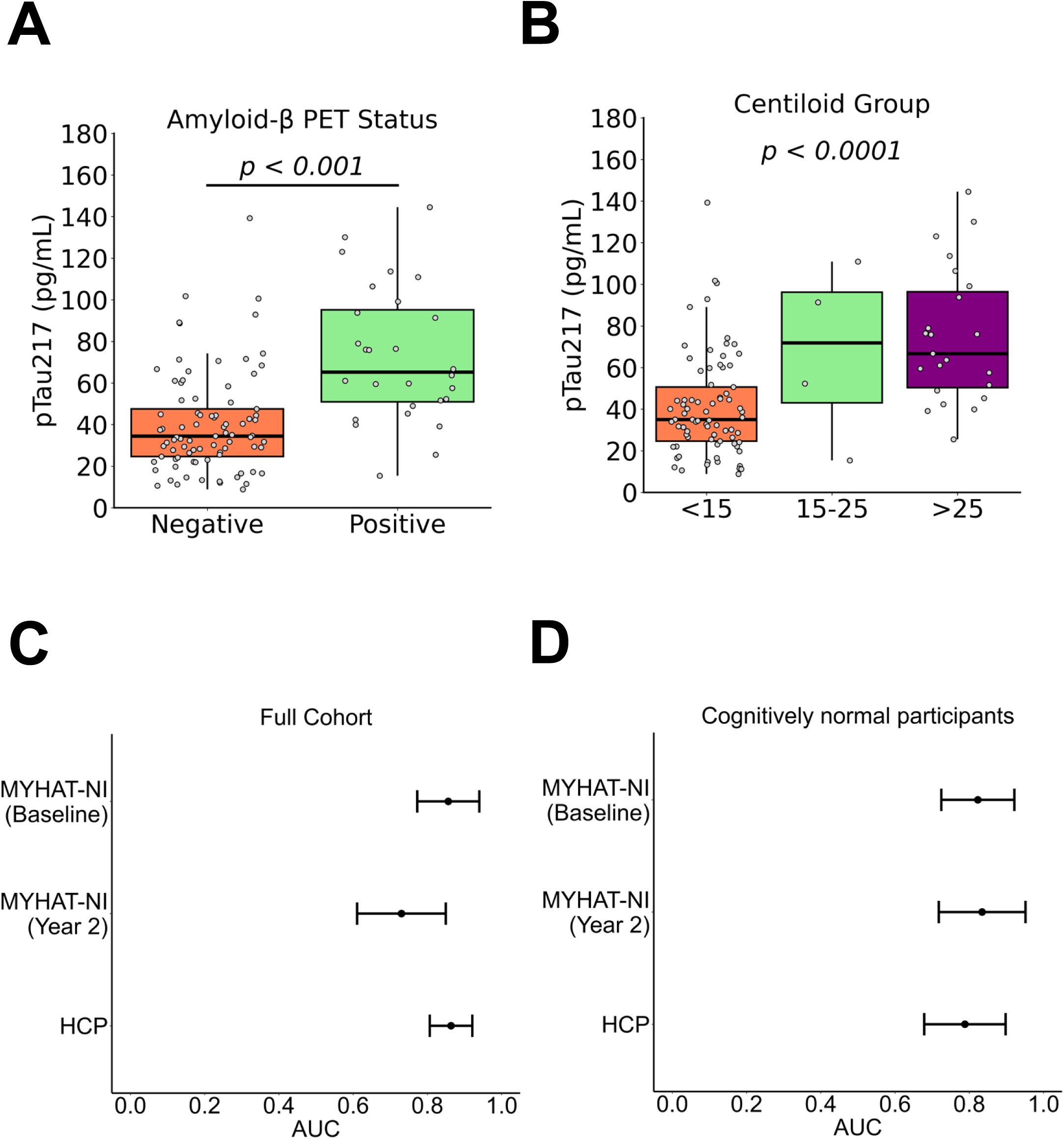
Diagnostic utility of the Pittsburgh p-tau217 assay in detecting. **A**β**-PET positivity.** (A) Distribution of plasma p-tau217 concentrations according to Aβ-PET status in the MYHAT-NI cohort at baseline. P-values were determined using the Wilcoxon Rank-Sum test. (B) Plasma p-tau217 levels in Aβ-negative (CL<15) to low-burden Aβ (CL15-25) and Aβ-positive (CL>25) in the MYHAT-NI cohort at baseline. P-values were determined from post-hoc tests following Kruskal-Wallis, with Bonferroni corrections to account for multiple comparisons. (C-D) AUC of Pittsburgh p-tau217 assay measurements for distinguishing Aβ-from Aβ+ participants across the full cohort (C), as well as in cognitively normal participants (D) in the MYHAT-NI and HCP cohorts.

In cognitive normal participants, plasma p-tau217 levels at baseline distinguished the groups with an AUC of 0.82 (95% CI = 0.72-0.92), and this separation was slightly better at the two-year visit with an AUC of 0.83 (95% CI = 0.72-0.95) (Figure 6D). Similarly, in the HCP cohort, the two groups were separated with an AUC of 0.79 (95% CI = 0.68-0.90) (Figure 6D).

We also evaluated the association between plasma p-tau217 levels and amyloid PET burden using a three-group classification system based on the Centiloid scale. There was a significant association between Aβ-PET centiloids (CLs) and plasma p-tau217 levels in the MYHAT-NI cohort (p < 0.001; Figure 6B) and the HCP cohort (p = 0.098; Supplemental Figure 5B). When plotted according to Aβ-PET CLs at baseline in the MYHAT-NI cohort, p-tau217 levels exhibited stepwise increases from the Aβ-PET-negative group (CL < 15) to the low-burden Aβ-PET group (CL 15-25), and to the Aβ-PET-positive group (CL > 25) (Figure 6B).

### 3.9 Association of Pitt p-tau217 levels with tau pathology

In the MYHAT-NI cohort, plasma p-tau217 levels were significantly elevated in tau-PET-positive compared with tau-PET-negative individuals at both baseline (1.4-fold; p = 0.014) ((Figure 7A) and 2-year visit (1.7-fold; p < 0.001) (data not shown). The discriminatory accuracy at baseline was only AUC 0.52 (95% CI = 0.40-0.65), which improved to 0.74 (95% CI = 0.61-0.87) at the two-year visit (Figure 7B). In individuals with normal cognition, plasma p-tau217 levels at baseline distinguished the groups with a poor AUC of 0.48 (95% CI = 0.36-0.60), but this separation was further strengthened at the two-year visit with an AUC of 0.79 (95% CI = 0.64-0.94) (Figure 7C).

**Figure 7.**
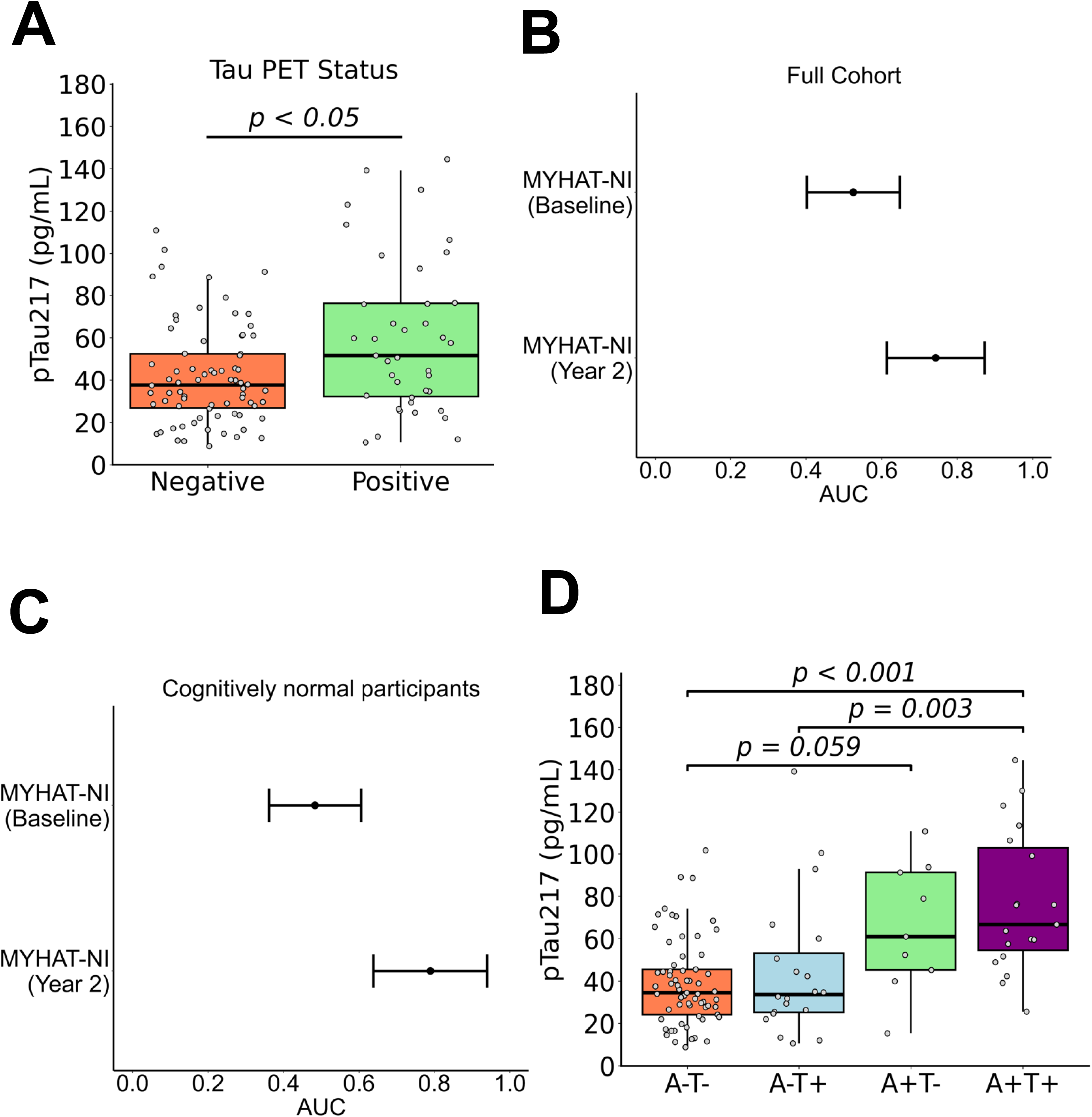
Diagnostic utility of the Pittsburgh p-tau217 assay for tau-PET positivity. (A) Distribution of plasma p-tau217 concentrations according to tau-PET status in the MYHAT-NI cohort at baseline. P value was determined using the Wilcoxon Rank-Sum test. (B) Plasma p-tau217 levels across AT groups at baseline in the MYHAT-NI cohort. P values were based on post-hoc tests following Kruskal-Wallis, with Bonferroni corrections. (C-D) AUC of Pittsburgh p-tau217 assay measurements for distinguishing tau PET-from tau PET+ participants across the full cohort (C), as well as in cognitively normal participants (D) in the MYHAT-NI cohort.

### 3.10 Plasma p-tau217 levels in relation to joint Aβ plaques and tau NFT status by PET

In MYHAT-NI at baseline, when categorized by concomitant Aβ and tau PET (A and T) status, plasma p-tau217 showed significant increases from A-T-to the A+T+ group (Figure 7D). The A+T+ group exhibited the highest levels compared with the A-T-, A+T-, and A-T+ groups (Figure 7B). The mean [SD] of p-tau217 (pg/mL) concentrations in MYHAT-NI at baseline were A−T−, 38.94 [20.65]; A−T+, 44.75 [33.02]; A+T−, 65.43 [30.59]; and A+T+, 76.85 [33.49], showing increasing levels when Aβ and tau pathologies are jointly abnormal.

## DISCUSSION

In this study, we have addressed a critical shortcoming in the literature regarding plasma p-tau217; namely if p-tau217 assays built using the same ultrasensitive technology but different antibodies result in similar performances. We achieved this by developing the novel plasma Pitt p-tau217 assay employing a new rabbit mAb that is distinct from the single mouse monoclonal antibody from ALZpath used in most p-tau217 immunoassays available. Our results demonstrate that the Pittsburgh p-tau217 assay signal in human plasma was elevated in antemortem samples from autopsy-verified sporadic AD and ADAD individuals. Moreover, the assay showed high accuracies in separating Aβ+ and Aβ-individuals across two cohorts including mostly cognitively normal older participants, as well as moderate accuracies in distinguishing tau-PET-positive and -negative individuals. Importantly, these results were strongly correlated with those obtained with the ALZpath p-tau217 assay, which uses the ALZpath antibody widely applied in most p-tau217 assays to date. These findings establish the utility of the Pittsburgh p-tau217 and underscores that p-tau217-specific assays established using Simoa technology can be highly accurate irrespective of if a commonly used antibody is used or if a novel antibody is applied.

With the exception of the Lumipulse and Janssen p-tau217 assays^33, 34^ that use different antibodies but account for a relatively small proportion of studies on p-tau217, all other research use only (RUO) and *in vitro* diagnostic immunoassay methods on p-tau217 currently use the ALZpath antibody. Hence, the literature is heavily skewed toward data obtained using this antibody, making it difficult to explain if the stellar performance on p-tau217 is either antibody-dependent or simply due to p-tau217 being an excellent biomarker. Our results support the latter conclusion, showing that novel p-tau217 antibodies can be used to develop high performance immunoassays. We chose to compare the Pittsburgh and ALZpath p-tau217 assays as like-for-like methods in the sense that: (1) they both use Simoa technology; (2) both assays use monoclonal antibodies that bind p-tau217 specifically without cross-reactivity to neighboring phosphorylation sites, as seen with the Janssen p-tau217+ assay for example; and (3) they both partner their p-tau217 antibody with an N-terminal phosphorylation-independent detection antibody.

The biochemical characterization of the novel p-tau217 antibody demonstrated high specificity to the p-tau217 site, even in the presence of phosphorylation at neighboring sites. In our immunohistochemistry study, the p-tau217 immunoreactivity observed in sections of middle temporal gyrus and in the hippocampus from advanced-stage AD brain identified morphologically diverse NFTs from pre-tangles to mature tangles, as seen in previous reports using p-tau217 and several other antibodies targeting tau phosphorylation at threonine epitopes^35–38^. In agreement with these studies, our results show that the p-tau217 antibody used in the current report identifies the full range of neurofibrillary pathology type including tangles, dystrophic neurites in neuritic plaques, and neuropil threads, further supporting the use of this antibody to detect tau pathology characteristic of AD that may be the source of the p-tau217 containing tau assayed in biological fluids. The robust immunostaining of dystrophic neurites in neuritic plaques may explain good correlation between plasma levels of p-tau217 and amyloid pathology detectable with PET, as noted in the current study and several other reports^36, 39^.

The Pittsburgh p-tau217 assay expands the repertoire of plasma p-tau217 immunoassay methods available on the market, particularly those from academic laboratories. At present, only a single blood-based p-tau217 immunoassay has been reported from an academic center^32^ with the rest all from commercial sources. RUO assays built from academic labs tend to be cheaper to implement, and amenable to experimental changes that can be performed to accelerate research inquiry without concerns about infringing on commercially driven intellectual properties. At a time where research funding is difficult to come by, having an in-house assay that will allow scientists to perform highly reliable p-tau217 testing at a fraction of the current costs should be advantageous. We estimate that the Pittsburgh p-tau217 assay costs a third of the ALZpath p-tau217 method, confirming its cost effectiveness. The strong correlations of the Pittsburgh and ALZpath p-tau217 assays show that they can both be used perhaps interchangeably to assess the pathophysiology of AD, without compromising on assay quality and classification accuracies.

Strengths of this study include the new p-tau217 antibody biochemical characterization, assay development, analytical and clinical validation. Moreover, clinical validation studies were performed in multiple cohorts including in one with matched antemortem plasma and postmortem neuropathological diagnosis, and others from population-based sources. Assay performance was also compared with that of an established one i.e., the ALZpath assay.

In conclusion, this study reports the biochemical characterization of a novel p-tau217 antibody and its use in developing an innovative Pittsburgh p-tau217 assay. This assay demonstrated equivalent performance with the widely used ALZpath p-tau217 immunoassay, which shares a similar design and is measured on the same Quanterix Simoa analytical platform as the new method. The Pittsburgh p-tau217 assay is an accurate sensitive, cost-effective and accessible research tool to investigating AD pathophysiology in research settings.

## Data Availability

All data produced in the present study are available upon reasonable request to the authors provided data sharing agreement is in place and measures to safeguard data integrity are ensured. Any request will first be reviewed to ensure that it does not infringe on any intellectual property and does not breach IRB approved procedures.

## Acknowledgements

We thank the Pitt ADRC, MYHAT and HCP study participants and their families and caregivers.

## Conflicts

TKK has consulted for Quanterix Corporation, SpearBio Inc., and Neurogen Biomarking LLC., has served on advisory board for Neurogen Biomarking LLC. (which comes with minority stock equity interest), and has received in-kind research support from Janssen Research Laboratories and Alamar Biosciences, outside the submitted work. He has received honoraria for speaker/grant review engagements from the NIH, UPENN, UW-Madison, Advent Health, Brain Health conference, Barcelona-Pittsburgh conference, the International Neuropsychological Society, the Icahn School of Medicine at Mount Sinai and CQDM Canada, all outside of the submitted work. TKK is an inventor on several patents and provisional patents regarding biofluid biomarker methods, targets and reagents/compositions, that may generate income for the institution and/or self should they be licensed and/or transferred to another organization. XZ, EEA and MDI are inventors on University of Pittsburgh provisional patents together with TKK. The other authors report no conflict of interest.

## Funding sources

The HCP study, MYHAT-NI study and the Pittsburgh ADRC are funded by NIH grants R01AG072641, R01 AG052521 and P30 AG066468. MDI and EEA were supported by NIH grants P01AG014449 and P01AG025204. TKK and the Karikari Laboratory were supported by the NIH (R01AG083874, U24AG082930, P30AG066468, RF1AG052525, R01AG053952, R37AG023651, RF1AG025516, R01AG073267, R01AG075336, R01AG072641, P01AG025204), The Cañizares Anderson Fund, and a professorial endowment from the Department of Psychiatry, University of Pittsburgh. The content is solely the responsibility of the authors and does not necessarily represent the official views of the National Institutes of Health or any other funder.

## Consent statement

All participants provided written consent, and the University of Pittsburgh Institutional Review Board approved the study.

## REFERENCES

1. Karikari TK, Ashton NJ, Brinkmalm G, et al. Blood phospho-tau in Alzheimer disease: analysis, interpretation, and clinical utility. Nat Rev Neurol 2022; 18: 400–418. 20220518. DOI: 10.1038/s41582-022-00665-2.

2. Gonzalez-Ortiz F, Kac PR, Brum WS, et al. Plasma phospho-tau in Alzheimer’s disease: towards diagnostic and therapeutic trial applications. Mol Neurodegener 2023; 18: 18. 20230316. DOI: 10.1186/s13024-023-00605-8.

3. Chong JR, Ashton NJ, Karikari TK, et al. Blood-based high sensitivity measurements of beta-amyloid and phosphorylated tau as biomarkers of Alzheimer’s disease: a focused review on recent advances. J Neurol Neurosurg Psychiatry 2021; 92: 1231–1241. 20210911. DOI: 10.1136/jnnp-2021-327370.

4. Karikari TK, Pascoal TA, Ashton NJ, et al. Blood phosphorylated tau 181 as a biomarker for Alzheimer’s disease: a diagnostic performance and prediction modelling study using data from four prospective cohorts. Lancet Neurol 2020; 19: 422–433. DOI: 10.1016/S1474-4422(20)30071-5.

5. Karikari TK, Benedet AL, Ashton NJ, et al. Diagnostic performance and prediction of clinical progression of plasma phospho-tau181 in the Alzheimer’s Disease Neuroimaging Initiative. Mol Psychiatry 2021; 26: 429–442. 20201026. DOI: 10.1038/s41380-020-00923-z.

6. Janelidze S, Mattsson N, Palmqvist S, et al. Plasma P-tau181 in Alzheimer’s disease: relationship to other biomarkers, differential diagnosis, neuropathology and longitudinal progression to Alzheimer’s dementia. Nat Med 2020; 26: 379–386. 20200302. DOI: 10.1038/s41591-020-0755-1.

7. Palmqvist S, Janelidze S, Quiroz YT, et al. Discriminative Accuracy of Plasma Phospho-tau217 for Alzheimer Disease vs Other Neurodegenerative Disorders. JAMA 2020; 324: 772–781. DOI: 10.1001/jama.2020.12134.

8. Ashton NJ, Pascoal TA, Karikari TK, et al. Plasma p-tau231: a new biomarker for incipient Alzheimer’s disease pathology. Acta Neuropathol 2021; 141: 709–724. 20210214. DOI: 10.1007/s00401-021-02275-6.

9. Kac PR, Gonzalez-Ortiz F, Emersic A, et al. Plasma p-tau212 antemortem diagnostic performance and prediction of autopsy verification of Alzheimer’s disease neuropathology. Nat Commun 2024; 15: 2615. 20240323. DOI: 10.1038/s41467-024-46876-7.

10. Zeng X, Lafferty TK, Sehrawat A, et al. Multi-analyte proteomic analysis identifies blood-based neuroinflammation, cerebrovascular and synaptic biomarkers in preclinical Alzheimer’s disease. Mol Neurodegener 2024; 19: 68. 20241010. DOI: 10.1186/s13024-024-00753-5.

11. Barthelemy NR, Salvado G, Schindler SE, et al. Highly accurate blood test for Alzheimer’s disease is similar or superior to clinical cerebrospinal fluid tests. Nat Med 2024; 30: 1085–1095. 20240221. DOI: 10.1038/s41591-024-02869-z.

12. Kac PR, Gonzalez-Ortiz F, Emersic A, et al. Plasma p-tau212: antemortem diagnostic performance and prediction of autopsy verification of Alzheimer’s disease neuropathology. medRxiv 2023 20231211. DOI: 10.1101/2023.12.11.23299806.

13. Ashton NJ, Janelidze S, Mattsson-Carlgren N, et al. Differential roles of Abeta42/40, p-tau231 and p-tau217 for Alzheimer’s trial selection and disease monitoring. Nat Med 2022; 28: 2555–2562. 20221201. DOI: 10.1038/s41591-022-02074-w.

14. Mila-Aloma M, Ashton NJ, Shekari M, et al. Plasma p-tau231 and p-tau217 as state markers of amyloid-beta pathology in preclinical Alzheimer’s disease. Nat Med 2022; 28: 1797–1801. 20220811. DOI: 10.1038/s41591-022-01925-w.

15. Ashton NJ, Puig-Pijoan A, Mila-Aloma M, et al. Plasma and CSF biomarkers in a memory clinic: Head-to-head comparison of phosphorylated tau immunoassays. Alzheimers Dement 2023; 19: 1913–1924. 20221112. DOI: 10.1002/alz.12841.

16. Mielke MM, Dage JL, Frank RD, et al. Performance of plasma phosphorylated tau 181 and 217 in the community. Nat Med 2022; 28: 1398–1405. 20220526. DOI: 10.1038/s41591-022-01822-2.

17. Brickman AM, Manly JJ, Honig LS, et al. Plasma p-tau181, p-tau217, and other blood-based Alzheimer’s disease biomarkers in a multi-ethnic, community study. Alzheimers Dement 2021; 17: 1353–1364. 20210213. DOI: 10.1002/alz.12301.

18. Xiao Z, Wu W, Ma X, et al. Plasma p-tau217, p-tau181, and NfL as early indicators of dementia risk in a community cohort: The Shanghai Aging Study. Alzheimers Dement (Amst) 2023; 15: e12514. 20231222. DOI: 10.1002/dad2.12514.

19. Aguillon D, Langella S, Chen Y, et al. Plasma p-tau217 predicts in vivo brain pathology and cognition in autosomal dominant Alzheimer’s disease. Alzheimers Dement 2023; 19: 2585–2594. 20221226. DOI: 10.1002/alz.12906.

20. Hyman BT and Trojanowski JQ. Consensus recommendations for the postmortem diagnosis of Alzheimer disease from the National Institute on Aging and the Reagan Institute Working Group on diagnostic criteria for the neuropathological assessment of Alzheimer disease. J Neuropathol Exp Neurol 1997; 56: 1095–1097. DOI: 10.1097/00005072-199710000-00002.

21. Braak H, Alafuzoff I, Arzberger T, et al. Staging of Alzheimer disease-associated neurofibrillary pathology using paraffin sections and immunocytochemistry. Acta Neuropathol 2006; 112: 389–404. 20060812. DOI: 10.1007/s00401-006-0127-z.

22. Ikonomovic MD, Abrahamson EE, Uz T, et al. Increased 5-lipoxygenase immunoreactivity in the hippocampus of patients with Alzheimer’s disease. J Histochem Cytochem 2008; 56: 1065–1073. 20080804. DOI: 10.1369/jhc.2008.951855.

23. Andreasson U, Perret-Liaudet A, van Waalwijk van Doorn LJ, et al. A Practical Guide to Immunoassay Method Validation. Front Neurol 2015; 6: 179. 20150819. DOI: 10.3389/fneur.2015.00179.

24. Mirra SS, Heyman A, McKeel D, et al. The Consortium to Establish a Registry for Alzheimer’s Disease (CERAD). Part II. Standardization of the neuropathologic assessment of Alzheimer’s disease. Neurology 1991; 41: 479–486. DOI: 10.1212/wnl.41.4.479.

25. Braak H and Braak E. Neuropathological stageing of Alzheimer-related changes. Acta Neuropathol 1991; 82: 239–259. DOI: 10.1007/BF00308809.

26. Thal DR, Rub U, Orantes M, et al. Phases of A beta-deposition in the human brain and its relevance for the development of AD. Neurology 2002; 58: 1791–1800. DOI: 10.1212/wnl.58.12.1791.

27. Lopez OL, Becker JT, Klunk W, et al. Research evaluation and diagnosis of probable Alzheimer’s disease over the last two decades: I. Neurology 2000; 55: 1854–1862. DOI: 10.1212/wnl.55.12.1854.

28. Lopez OL, Becker JT, Chang Y, et al. Amyloid deposition and brain structure as long-term predictors of MCI, dementia, and mortality. Neurology 2018; 90: e1920–e1928. 20180425. DOI: 10.1212/WNL.0000000000005549.

29. Snitz BE, Tudorascu DL, Yu Z, et al. Associations between NIH Toolbox Cognition Battery and in vivo brain amyloid and tau pathology in non-demented older adults. Alzheimers Dement (Amst) 2020; 12: e12018. 20200515. DOI: 10.1002/dad2.12018.

30. Royse SK, Minhas DS, Lopresti BJ, et al. Validation of amyloid PET positivity thresholds in centiloids: a multisite PET study approach. Alzheimers Res Ther 2021; 13: 99. 20210510. DOI: 10.1186/s13195-021-00836-1.

31. LeDell E, van der Laan MJ and Petersen M. AUC-Maximizing Ensembles through Metalearning. Int J Biostat 2016; 12: 203–218. DOI: 10.1515/ijb-2015-0035.

32. Gonzalez-Ortiz F, Ferreira PCL, Gonzalez-Escalante A, et al. A novel ultrasensitive assay for plasma p-tau217: Performance in individuals with subjective cognitive decline and early Alzheimer’s disease. Alzheimers Dement 2024; 20: 1239–1249. 20231117. DOI: 10.1002/alz.13525.

33. Van Hulle C, Jonaitis EM, Betthauser TJ, et al. An examination of a novel multipanel of CSF biomarkers in the Alzheimer’s disease clinical and pathological continuum. Alzheimers Dement 2021; 17: 431–445. 20201218. DOI: 10.1002/alz.12204.

34. Vandijck M, Dekeyser F, Lambrechts C, et al. Preliminary performance of the Lumipulse G pTau 217 Plasma prototype assay on plasma samples. Alzheimer’s & Dementia 2023; 19: e079647.

35. Moloney CM, Labuzan SA, Crook JE, et al. Phosphorylated tau sites that are elevated in Alzheimer’s disease fluid biomarkers are visualized in early neurofibrillary tangle maturity levels in the post mortem brain. Alzheimers Dement 2023; 19: 1029–1040. 20220803. DOI: 10.1002/alz.12749.

36. Wennstrom M, Janelidze S, Nilsson KPR, et al. Cellular localization of p-tau217 in brain and its association with p-tau217 plasma levels. Acta Neuropathol Commun 2022; 10: 3. 20220106. DOI: 10.1186/s40478-021-01307-2.

37. Hanes J, Kovac A, Kvartsberg H, et al. Evaluation of a novel immunoassay to detect p-tau Thr217 in the CSF to distinguish Alzheimer disease from other dementias. Neurology 2020; 95: e3026–e3035. 20200924. DOI: 10.1212/WNL.0000000000010814.

38. Bellier JP, Cai Y, Alam SM, et al. Uncovering elevated tau TPP motif phosphorylation in the brain of Alzheimer’s disease patients. Alzheimers Dement 2024; 20: 1573–1585. 20231202. DOI: 10.1002/alz.13557.

39. Mattsson-Carlgren N, Janelidze S, Bateman RJ, et al. Soluble P-tau217 reflects amyloid and tau pathology and mediates the association of amyloid with tau. EMBO Mol Med 2021; 13: e14022. 20210505. DOI: 10.15252/emmm.202114022.

