## Supplemental Figure 1 for "Pittsburgh plasma p-tau217: classification accuracies for autosomal dominant and sporadic Alzheimer’s disease in the community"

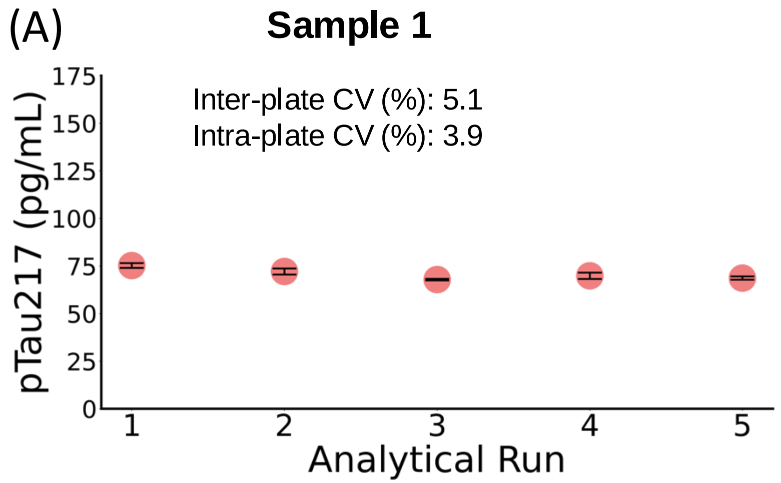


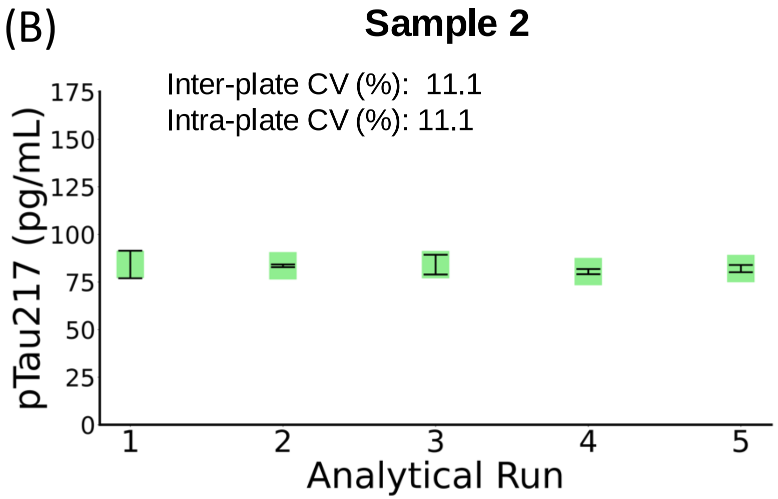


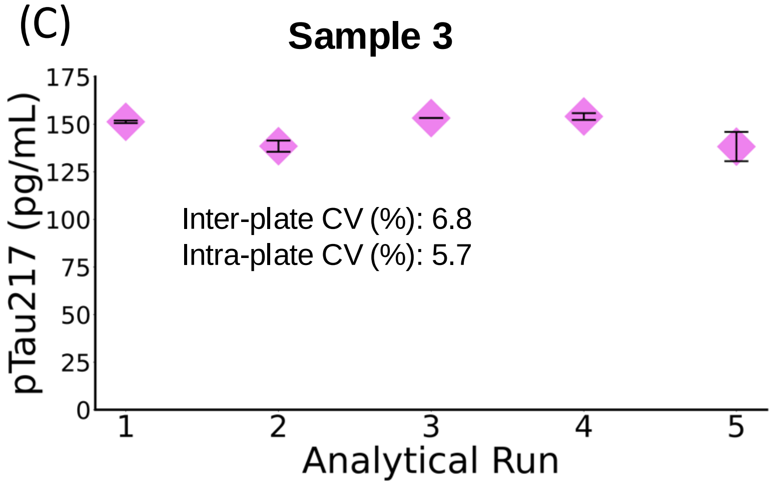


(D)

| | | Cohort | |
|---|---|---|---|
| | | MYHAT-NI | PSEN1 |
| Inter-plate CV (%) | Sample 1 | 4 | N/A |
| | Sample 2 | 5.8 | N/A |
| | Sample 3 | 16.8 | N/A |
| Intra-plate CV (%) | Sample 1 | 3.6 | 12.2 |
| | Sample 2 | 2.1 | 18.8 |
| | Sample 3 | 5.7 | 13 |

**Supplemental Figure 1. Precision Assessment of the Pittsburgh p-tau217 assay.** (A-C) Precision assessment based on results from three pooled plasma samples measured across five independent runs, with duplicate measurements per run. Data points represent the mean concentration of duplicate measurements for each run, and error bars indicate the standard error of the mean (SEM). (D) Precision evaluation of the same pooled plasma samples analyzed during the cohort sample analysis.
