## Supplemental Figure 2 for "Pittsburgh plasma p-tau217: classification accuracies for autosomal dominant and sporadic Alzheimer’s disease in the community"

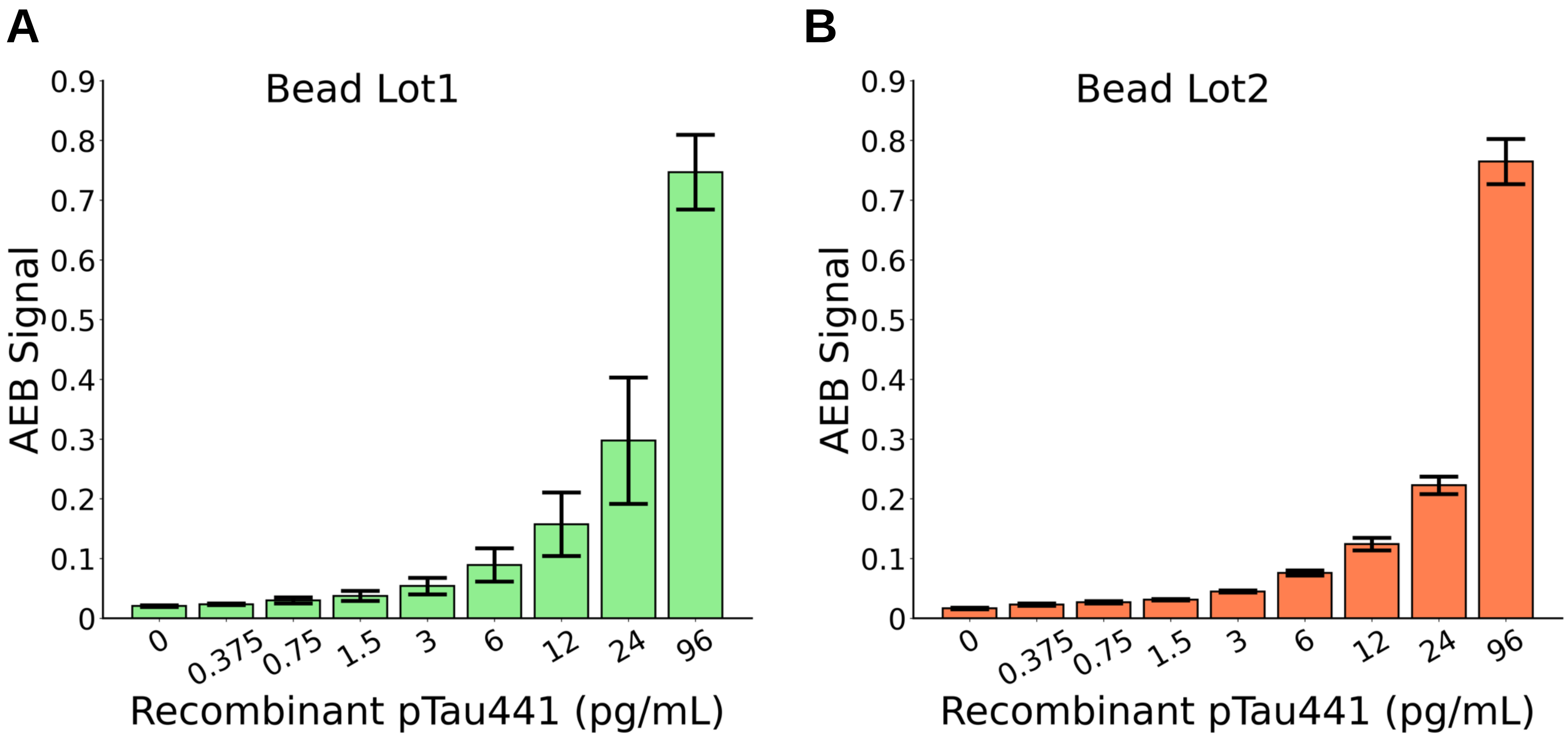


**Supplemental Figure 2. Cross validation of the Pittsburgh p-tau217 assay.** The impact of antibody lots were evaluated by running standard curves (p-tau441; 0.375–96pg/ml) using antibodies from two different lots. Consistent signals were detected at every data point, indicating the high precision of the Pittsburgh p-tau217 assay regardless of the antibody lots or instruments used. Data points represent mean concentration of in duplicates in two independent experiment runs and error bars represent ± SEM. Samples were run in duplicates.
