## Supplemental Figure 3 for "Pittsburgh plasma p-tau217: classification accuracies for autosomal dominant and sporadic Alzheimer’s disease in the community"

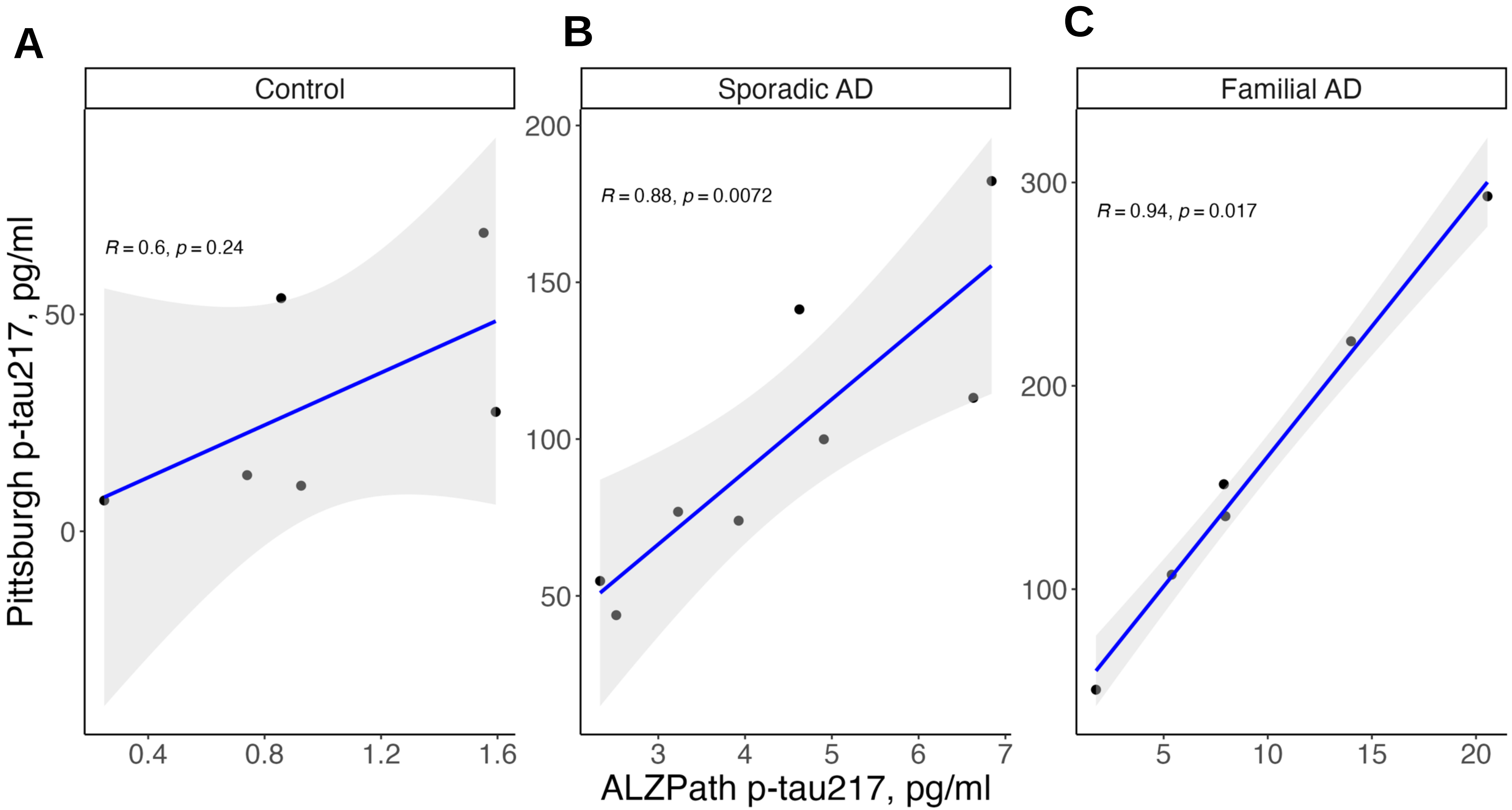


**Supplemental Figure 3. Performance comparison of the Pittsburgh vs. ALZpath p-tau217 assays to distinguish sporadic and familial AD from non-AD controls.** Scatterplot distributions illustrating the correlation of p-tau217 levels measured using Pitt and ALZpath method in non-Alzheimer's disease controls and sporadic & familial Alzheimer's disease cases. ρ and p-values were determined using Spearman rank-based correlation. Blue lines indicated the least square regression lines.
