## Supplemental Figure 4 for "Pittsburgh plasma p-tau217: classification accuracies for autosomal dominant and sporadic Alzheimer’s disease in the community"

**A** **Full Cohort**

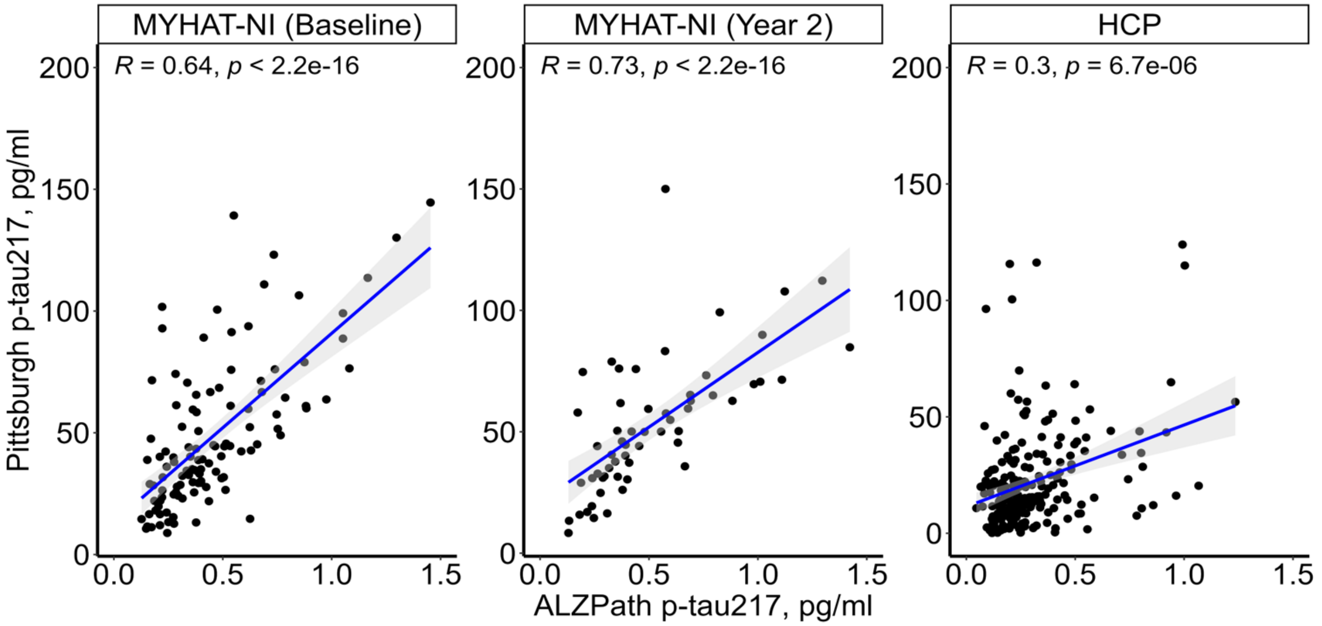


**B** **Cognitively normal participants**

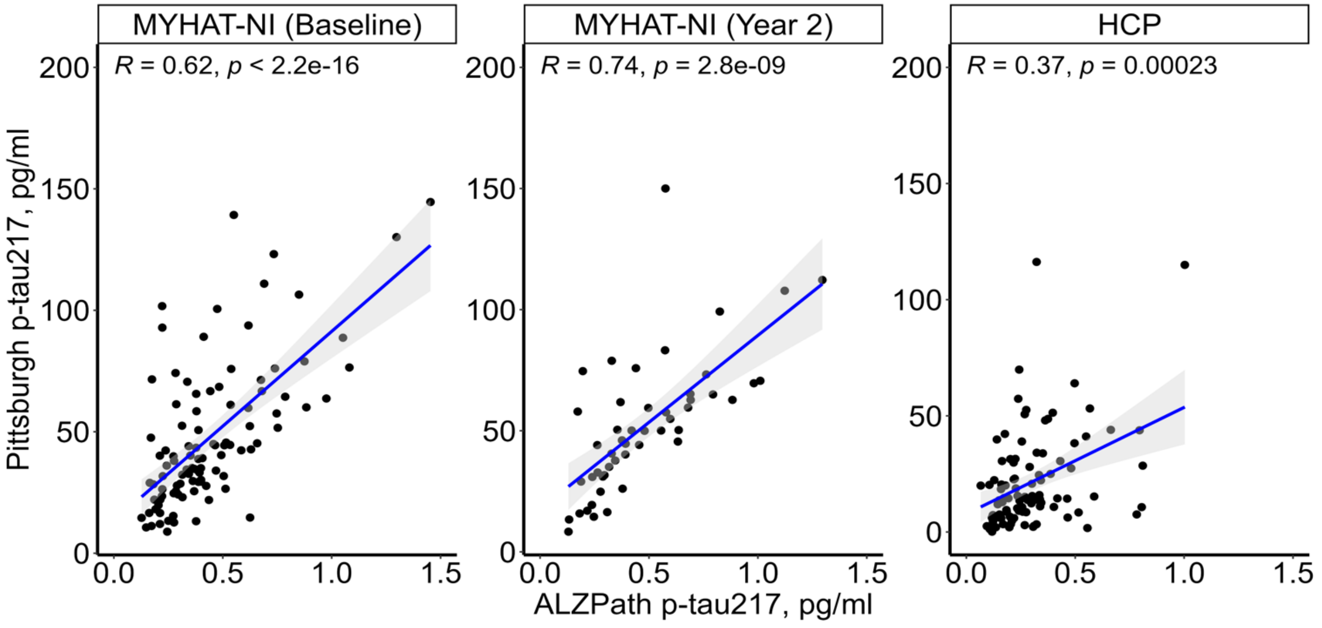


**Supplemental Figure 4. Spearman's correlation between p-tau217 measured by the Pittsburgh and ALZpath assays.** Scatterplots illustrating the correlation between Pittsburgh (y-axis) and ALZpath p-tau217 (x-axis) assays across full (A) and in cognitively normal participants (B) in MYHAT-NI cohort at baseline, and at a visit 2 years later and in HCP cohort. Blue lines indicated the least square regression lines. P-values are from significance testing of Spearman correlation
