## Supplemental Figure 5 for "Pittsburgh plasma p-tau217: classification accuracies for autosomal dominant and sporadic Alzheimer’s disease in the community"

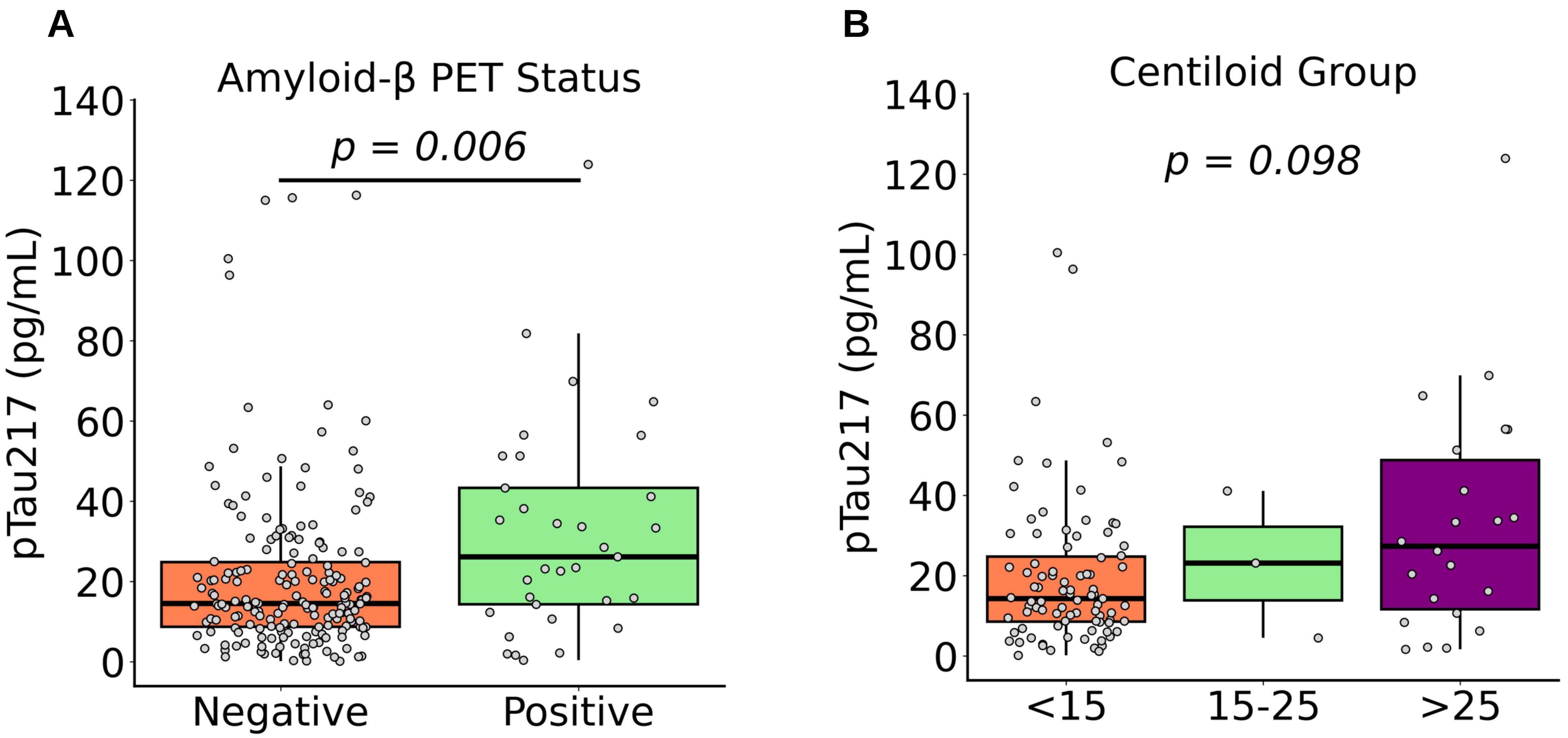


**Supplemental Figure 5. Classification utility of the Pittsburgh p-tau217 assay in a racially diverse HCP cohort.** (A) Distribution of plasma p-tau217 concentrations according to Aβ-PET status in the HCP cohort. P-values are from Wilcoxon Rank-Sum Test. (B) Plasma p-tau217 levels in Aβ-negative (CL<15) to low-burden Aβ (CL15-25) and Aβ-positive (CL>25) in the HCP cohort. P-values were computed using the Kruskal-Wallis test with Bonferroni correction for multiple comparisons.
